# Indoor Air Quality Intervention in Schools; Effectiveness of a Portable HEPA Filter Deployment in Five Schools Impacted by Roadway and Aircraft Pollution Sources

**DOI:** 10.1101/2022.01.12.22269175

**Authors:** Nancy Carmona, Edmund Seto, Timothy Gould, Jeffry H. Shirai, B.J. Cummings, Lisa Hayward, Timothy Larson, Elena Austin

**Affiliations:** Department of Environmental & Occupational Health Sciences, University of Washington, Seattle, WA 98195, USA; Department of Civil & Environmental Engineering, University of Washington, Seattle, WA 98195, USA

## Abstract

The Healthy Air, Healthy Schools Study was established in January 2020 to better understand the impact of ultrafine particles (UFP) on indoor air quality in communities surrounding Seattle-Tacoma (Sea-Tac) International Airport. The study team took multipollutant measurements indoor and outdoor air pollution at five participating school locations to infiltration indoors. The schools participating in this project were located within a 7-mile radius of Sea-Tac Airport and within 0.5 miles of an active flight path. Based on experimental measures in an unoccupied classroom, infiltration rates of a) Ultrafine particles of aircraft origin b) Ultrafine particles of traffic origin and c) Wildfire smoke or other outdoor pollutants were characterized before and after the introduction of a classroom based portable HEPA filter intervention. The portable HEPA cleaners were an effective short-term intervention to improve the air quality in classroom environments, reducing the ultrafine particles to approximately 1/10th of that measured outside. Before the HEPA filter deployment, approximately one-half of all outdoor UFPs were measured indoors. This study is unique in focusing on UFP in school settings and demonstrating through multivariate methods that the UFP measured in the classroom space is primarily of outdoor origin. Although existing research suggests that improvements to indoor air quality in homes can significantly improve asthma outcomes, further research is necessary to establish the benefit to student health and academic performance of improved air quality in schools.

## Background

Increasing evidence has highlighted the impacts of traffic-related outdoor air pollutants, including ultrafine particles, on communities living in proximity to aircraft descent paths within the United States and internationally. The recently completed MOV-UP study in King County, Washington, identified a clear, aircraft-associated footprint of ultrafine particles under flight paths. Monitoring campaigns conducted in communities near airports in Seattle,^2–4^ Los Angeles,^5–8^, Atlanta,^9^ Boston,^10^ New York^11^ and Amsterdam^12^ have all identified elevated levels of total UFP in proximity to international airports. This work has also highlighted differences in the pollutant mixtures between aircraft and roadway traffic sources,^6,11,13,14^ as well as differences in fuel-based emissions of UFP from aircraft and roadway traffic sources.^2,7^ Evidence is emerging that exposure to aircraft emissions is associated with negative health impacts. A recent 10-year retrospective population-based study in Los Angeles found a significant increase in pre-term births in women exposed to aircraft-related pollution during gestation, and this effect was found to be independent of the effect of roadway traffic pollution.^15^ A subsequent analysis of the same airport-impacted area near LAX established an increased risk of malignant brain cancer in individuals exposed to aircraft UFP.^16^ This, as well as previous work demonstrating short-term increases in inflammation in adults exposed to community air pollution in aircraft-impacted locations,^17^ suggests the need to implement measures to increase resilience in communities and establish long-term monitoring.

Resiliency in a community is improved when vulnerable members are provided with interventions designed to mitigate or remove their sources of exposure. School settings have been identified as priority environments for intervention and identification of exposure reduction strategies, particularly in response to extreme events such as wildfires.^18^ Currently, it is not well understood how UFP from outdoor sources may infiltrate into indoor environments. Experimental and theoretical simulations of particle movement suggest that the ability of particles to enter an indoor space from the outdoor air (infiltration) varies widely and depends on building characteristics such as ventilation system type, leaking through cracks and other openings as well as open windows.^19^ Possible infiltration factors for UFP range greatly across studies and building types and typically range from 10-70% infiltration.^19,20^ Important determining variables include a) building type; b) ventilation system parameters, including central vs. local units, filter type and manufacturer; and c) building management strategies.

Existing literature supports the notion that in-class performance of students is directly impacted by the air pollution level at their school. In Los Angeles, researchers studied how changes in ambient air pollution concentrations affected the performance of second-through sixth-grade students on standardized tests between 2002 and 2008.^21^ Comparisons were made between different cohorts within the same school to account for differences between schools, including differences in outdoor pollution, socioeconomic status of students and other factors that vary between schools. Researchers found that lower concentrations of daily outdoor particulate matter significantly increased mathematics and reading test scores. Similar associations between test scores and short-term air pollution concentrations have been observed nationally and internationally.

Previous efforts to evaluate the impact of interventions to remove air pollutants in indoor spaces are limited and generally focused on residential environments. A researcher in Texas examined the effect of rolling indoor air quality (IAQ) improvements at nearly every school in a single school district.^22^ This quasi-natural experiment indicated that student performance on standardized tests significantly improved following improvements in IAQ. Similarly, preliminary results from another quasi-natural experiment in California, where High Efficiency Particulate Air (HEPA) filters were installed in every classroom, office and common area for all schools within five miles of a potential gas leak (but not beyond), found that air filter usage led to a significant increase in mathematics and English scores, with test score improvements persisting into the following year.^23^

### Outdoor air pollution

Outdoor ambient air pollution, including fine particulate matter, has been demonstrated to have respiratory and cardiovascular effects^24^, impact birth weight and infant mortality^25,26^ and is hypothesized to be associated with the development of Alzheimer’s disease^27,28,29^. Regulatory monitoring does not capture exposure variation within communities particularly well; within-community exposures can be impacted by local sources such as highways, major roadways, construction or industrial facilities.

Exposures may also be disproportionately high in black and Hispanic minorities, which experience more air pollution than non-Hispanic whites^30^. Community-engaged research provides an opportunity for the public to learn about neighborhood-level exposures to ambient air pollution, health effects and mitigation strategies^31,32^.

In the United States, fine particulate matter with an aerodynamic diameter of ≤ 2.5 micrometers (µm) (PM_2.5_) is regulated by the Environmental Protection Agency (EPA) under the National Ambient Air Quality Standards (NAAQS)^26^. The EPA has affirmed a causal relationship between PM_2.5_ exposure and respiratory and cardiovascular effects, including the exacerbation of respiratory and cardiovascular disease^33,34^, as well as low birth weight and infant mortality from respiratory issues^25,26^. There are also new emerging health effects associated with exposure to air pollution and increased risk of cognitive function and decline in the vulnerable population of older adults^35,36^. While the Puget Sound region has lower levels of pollution than other large cities, health effects can occur in areas with low ambient air pollution^37,38^. Puget Sound’s high population density makes this an important public health concern, particularly for sensitive populations such as children and older adults.

There are various factors that affect the variability of PM_2.5_ over the Puget Sound region. The distribution of emissions affecting the Puget Sound airshed results from multiple sectors, including natural sources, vehicles, industries with combustion units, wood stoves and fireplaces^39^. Air pollution also varies by time of year depending on weather conditions and seasonal sources such as wood stoves^40^. Historically, days with the highest PM_2.5_ concentrations have overwhelmingly occurred during winter when wood is used as a heating source in fireplaces^41^. The second largest contributors of particulate matter are cars and trucks. Transportation sources including freeways, highways and major arterial roadways, railway, aircraft and marine vehicles contribute to regional and localized pollutant concentrations^42^.

Increasingly, wildfires are becoming a significant source of PM_2.5_ in the summers. In 2017, the wildfire season in the Puget Sound region resulted in 16 wildfire-impacted days, meaning 4.4% of days that year were “unhealthy for sensitive groups or unhealthy” according to the air quality index (AQI)^43^. Climate change can affect fire frequency, extent and severity,^44^ with models for California predicting that further climate change will amplify the duration of extreme fire weather by the end of the century^45,44^. Notably, exposure to wildfire PM_2.5_ has been found to be associated with cardiorespiratory mortality^46^. Public health officials recommend staying indoors to reduce wildfire PM_2.5_ exposures, but smoke infiltration may result in poor indoor air quality, which will be discussed later^47^.

Another air pollution source of particular interest is air transportation. Aircraft pollution sources are attracting more attention in the context of public health due to their UFP emissions. UFPs have an aerodynamic diameter less than 0.1 μm. Unlike for PM_2.5_, there is no EPA regulatory standard for UFPs. In the Puget Sound region, Sea-Tac International Airport lies about 13 miles (∼21 km) from downtown Seattle, but several smaller cities such as SeaTac, Burien, Des Moines and Normandy Park surround the airport. UFP levels have been found to be elevated near large airports and have different composition and size than those from road traffic.^48^ A study near Logan International Airport in Boston found that aviation emissions can significantly impact ambient residential areas up to 7.3 km from the airport^49^. In addition, a study near Schiphol International Airport in Amsterdam found short-term exposure to high levels of UFP to be associated with decreased lung function and cardiac function in young healthy adults^48^. Downwind from Los Angeles International Airport, concentrations of UFPs have been found four to five times the background concentration up to 10 km in distance^17^. Given the potential health effects related to proximity to airports, assessing UFP exposures attributed to Sea-Tac International Airport is vital for improving health disparities in the Puget Sound region.

### Indoor air pollution

Given that people spend 85% to 90% of their time indoors, the quality of indoor air is likely to have a significant impact on health, even though it is outdoor air that is regulated^50^. EPA exposure studies indicate that indoor levels of pollutants may be two to five times, and occasionally more than 100 times, higher than outdoor levels^51^. Some indoor gaseous air pollutants can be emitted by materials in the building, furniture finishes, paints, adhesive and particle board^52^. Indoor sources of particles include gas-fired appliances, cooking, vacuuming/cleaning activities and the generation of particles by people. Symptoms related to indoor air pollution include headaches, fatigue, shortness of breath, sinus congestion, coughing, sneezing, dizziness, nausea and irritation of the eye, nose, throat and skin^51^. There is substantial evidence that indoor environmental exposures to allergens (such as dust mites, pests and molds) play a role in triggering asthma symptoms^51^. These high exposures are concerning for sensitive populations, such as children. Nearly 1 in 13 school-aged children has asthma, which is the leading cause of school absenteeism due to chronic illness^51^. Indoor air pollution can also be detrimental to students’ academic performance^51^.

Children are thought to be especially vulnerable to exposure to UFP. Previous studies have confirmed that concentrations of UFP within schools is associated with indoor factors such as the number of students, the type of furnishings and window types. However, it is also inversely correlated with indoor CO_2_, suggesting that there is an important outdoor source of UFP in school buildings.^53^ A European review of UFP exposures of children suggests that the greatest predictors of high exposure in children were proximity to heavy traffic or near cooking and cleaning activities.^54^

Indoor air quality is directly impacted by the infiltration of ambient air pollution as particles move from the outdoors to indoors. Buildings are typically ventilated using mechanical and natural ventilation, transporting outdoor particles to the indoor environment^55^. Air can also enter buildings through openings, joints and cracks in walls, floors and ceilings and around windows and doors. The ventilation rate is equal to the number of times the air in an indoor space is completely replaced by outdoor air (outdoor air exchange rate or AER)^52^. Typical values of AER range from 0.25 or lower for a tightly sealed, efficient building to 1 or higher for leaky buildings^52^. Figure 1 illustrates the movement of particles into and out of an indoor environment.

**Figure 1.**
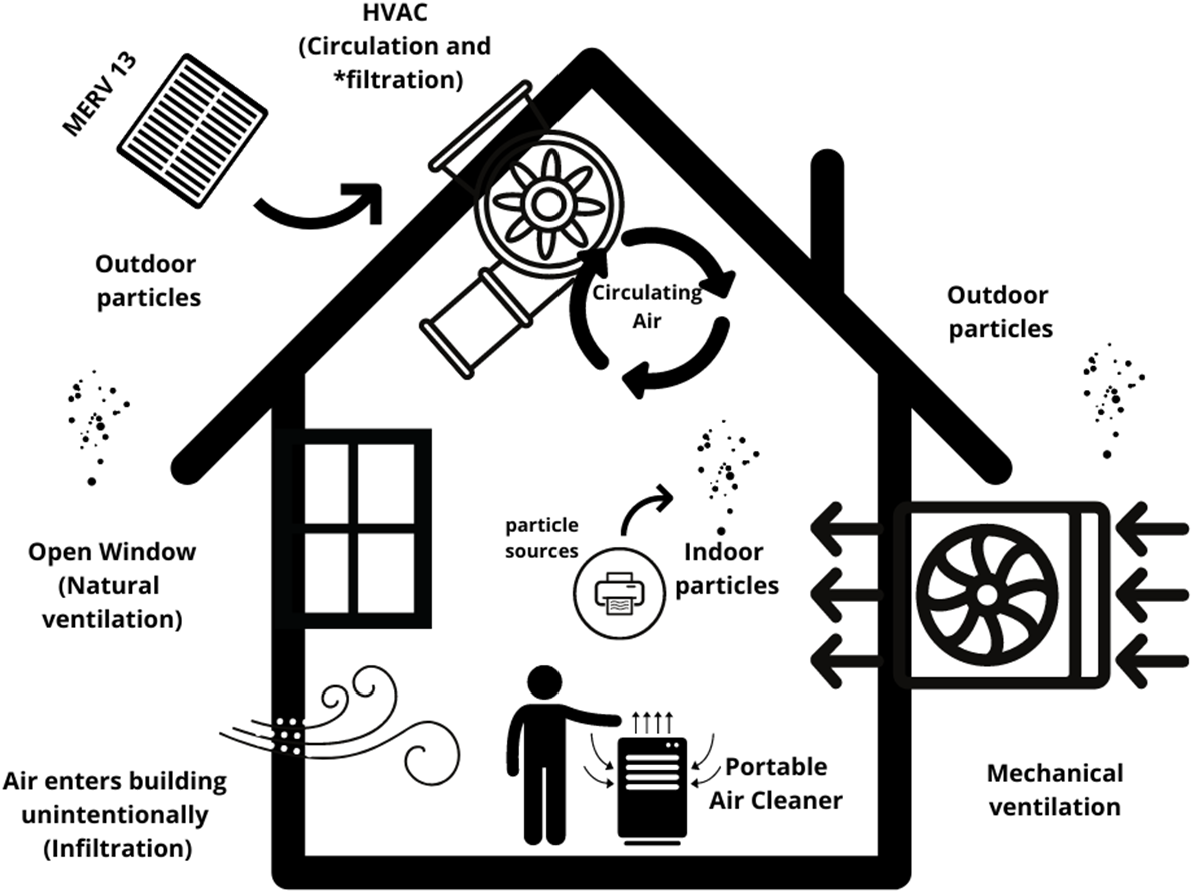
Indoor Air Pollution Dynamics

One strategy to improve indoor air quality is to upgrade the heating, ventilation and/or air conditioning (HVAC) system to filter out particles within a building^47^. A minimum efficiency reporting value (MERV) 13-rated filter or the highest rated filter the HVAC system can handle should be used for optimal particle reduction^47^. One limitation in filtration is that HVAC systems in older buildings may not be able to overcome the increased resistance to air flow of the higher efficiency filters. Portable air cleaners with HEPA filters can also improve indoor air quality by removing particulates from the air^47^. A HEPA filter, as defined by the EPA, is a type of pleated mechanical air filter that can remove at least 99.97% of dust, pollen, mold, bacteria and any airborne particles with a size of 0.3 microns^56^. However, particles larger or smaller than 0.3 microns are trapped with a higher efficiency by all filter types^56^. HEPA filters have a MERV rating greater than 16^57^. Figure 2 shows the relationship between particle diameter and removal efficiency for various MERV ratings and HEPA filters.

**Figure 2.**
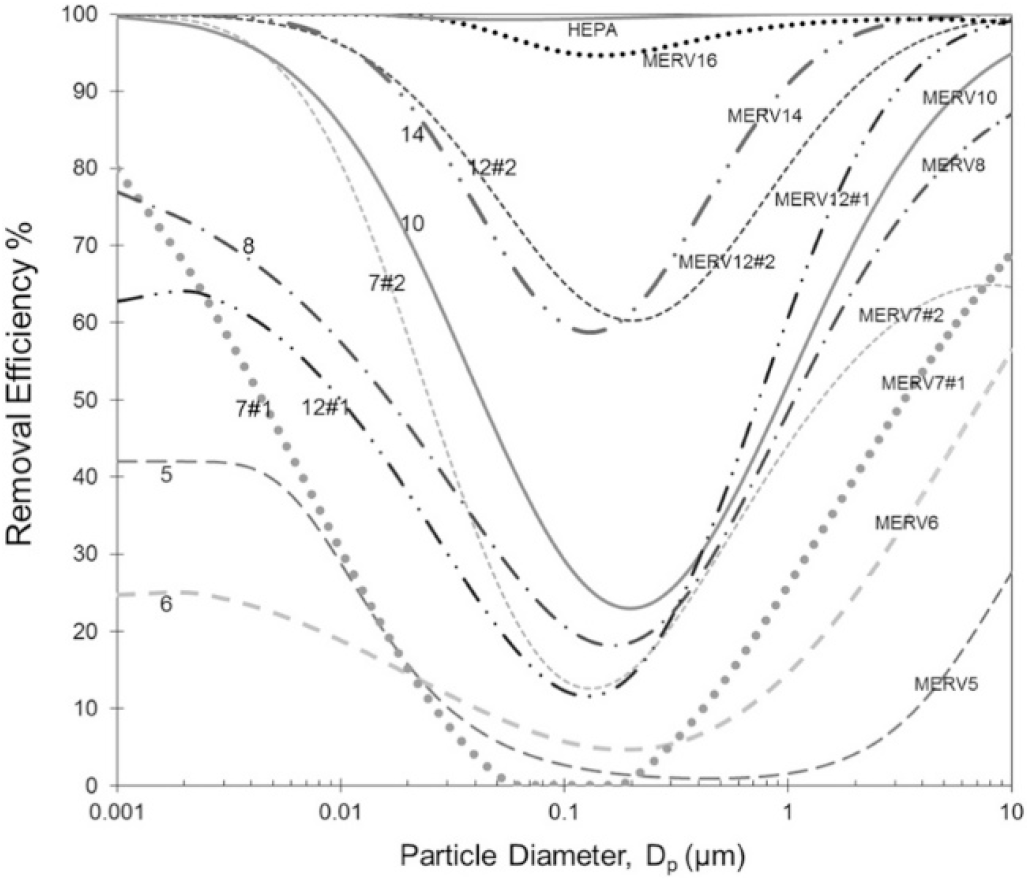
Particle size specific MERV filter efficiency. Source: Azimi et al. 2014

Building HVAC systems are typically characterized based on the MERV rating. The higher the rating, the better the filter is at removing specific sizes of particles^56^. The MERV rating is derived from a test method developed by the American Society of Heating, Refrigerating, and Air Conditioning Engineers (ASHRAE)^56^.

## METHODS

### Study area

The Healthy Air, Healthy Schools Study is being conducted in two phases. For Phase 1, the specific objectives were to 1) inform schools, districts and legislators on current building ventilation systems efficiency in removing outdoor particles, 2) quantify the current ability of ventilation solutions to remove indoor-generated particles; 3) identify any additional benefits and costs of in-room filtration and air handling interventions; 4) describe the infiltration rates of ultrafine particles of aircraft origin, ultrafine particles of traffic origin and wildfire smoke; and 5) communicate the results to all study partners.

Monitoring sites within the Federal Way and Highline school districts were selected by the University of Washington research team with guidance from the Federal Way and Highline Public Schools partners. Five schools in the Sea-Tac Airport flight path were selected to evaluate the impact of airport traffic. The participating school buildings represent a variety of air handling designs and building ages. Infiltration of outdoor air pollution into classroom spaces was measured under normal operating conditions and after deploying HEPA filters as a classroom-level intervention. Air monitoring took take place in spring and summer of 2021.

Phase 2 of the study is being conducted in FY 2021-2023. In Phase 2, the project will conduct a longer-term evaluation of the effect of air filtration on indoor air quality in 20 schools, characterize outdoor exposures at these school sites and evaluate the added benefit of the HEPA unit deployment.

### Portable air cleaner intervention

The portable air cleaner proposed for use in this study is the Blueair model 605 portable air cleaner with a HEPA-rated filter. Each device is evaluated by the Association of Home Appliance Manufacturers (AHAM) Institute to ensure it can provide a clean air delivery rate (CADR) for smoke and dust and supply adequate filtration for large spaces (∼800 square feet). The noise level is rated to be between 33 and 62 dB(A), depending on the fan speed setting. This device is expected to provide adequate filtration over a six-month period and a HEPA filter with 99.9% efficiency at capturing ultrafine and fine particles, including those originating from wildland fire smoke.

### School sites

Sites were selected in consultation with school district partners to represent a range of building ages as well as proximity to flight paths and roadway traffic. All school sites selected were within 0.5 miles of an active flight path serving Sea-Tac Airport and within a 7-mile radius of the airport. The classrooms where monitoring occurred were selected by school staff to be representative of the school or a particular part of the building, as well as rooms that were vacant of students at the time of our sampling. Characteristics of the monitored classrooms are shown in Table 1.

**Table 1.** School classroom dimension and volume

| School | Distance From Airport | Room Area, full dimensions (ft)<sup>a</sup> | Ceiling Height (ft) | Room Volume (ft<sup>3</sup>) | Room Volume (m<sup>3</sup>) |
| --- | --- | --- | --- | --- | --- |
| School A 1<sup>st</sup> flr. | 1.5 miles | 30.2 x 29.2 | 9.9 | 8,725.3 | 247.1 |
| **School A 2<sup>nd</sup> flr.** | 1.5 miles | 35.0 x 28.0 | 9.9 | 9,227.5 | 261.3 |
| **School B** | 2.1 miles | 32.0 x 28.7 | 9.3 to 12.9<sup>b</sup> | 10,053.7 | 284.7 |
| **School C** | 7.2 miles | 31.8 x 26.5 | 10.4 | 8,750.3 | 247.8 |
| **School D** | 0.5 miles | 31.7 x 23.1 | 9.1 | 6,574.7 | 186.2 |
| **School E** | 5.3 miles | 32.0 x 30.0 | 8.2 | 7,840 | 222.0 |

<sup>a</sup> Some classrooms have walled-off corner sections so are not fully rectangular
<sup>b</sup> Sloping ceiling, minimum height near windows and maximum by interior hallway

### Outdoor air exchange rate

The outdoor air exchange rate (AER) was measured at each site visit. This measure of air exchange reflects the exchange of air between the indoor and outdoor space and is a component of total air exchange rate that also includes recirculation through the HVAC filter system. Since our primary interest was in the movement of outdoor air into the indoor space, we focused on the measure of outdoor AER in this project. We considered three different models to calculate the outdoor AER. AER measurements were conducted using a protocol developed by the Harvard University T.H. Chan School of Public Health’s Healthy Buildings Program.^58^ Since we were able to conduct our measurements in unoccupied classrooms, we used the CO_2_ decay method to determine the air exchange rate from among the options presented in the Harvard Healthy Buildings Program guide.

The method involves elevating the CO_2_ concentration in the test classroom and then measuring the declining CO_2_ concentration over time to enable determination of the decay rate. Dry ice was used as the source of CO_2_ to elevate the inside concentration. A tray was filled with dry ice and two box fans operated in the room to thoroughly mix the CO_2_ as the concentration increased. With CO_2_ elevated to four times or more the background level, the dry ice was removed from the room and the mixing fans shut off to begin the decay of CO_2_ concentration while the field technician exited the classroom.

Two CO_2_ analyzers were used to characterize the CO_2_ concentration, one inlet near the center of the room and the other close to the windows along an outside wall of the classroom. Uniformity of the CO_2_ concentration within the room could be tracked over time, and with equivalent or very similar levels determined from the two monitors, we could then average the concurrent results as being representative for the entire room. The rate of decay without CO_2_ sources in the room is based on air exchange from (1) infiltration of air from the outside, and (2) the active ventilation system in the building. An adequate time series of CO_2_ decay is attained once the concentration drops to about one-third of the starting elevated level. The CO_2_ data from the time at which sources are removed and the decline begins through the time at which ventilation characteristics are altered by opening doors or people reentering the room will define the decay rate of CO_2_ used to determine the air exchange rate. The measured in-room CO_2_ less the ambient outdoor CO_2_ concentration is the quantity of interest to use in determining the air exchange rate.

Three models were considered to quantify the decay rate observed during our measurement of CO_2._ The first model we considered was an exponential decay model:

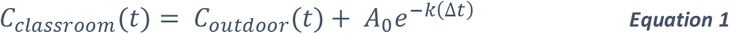

where *C*_*classroom*_ is the concentration within the classroom; C_outdoor_(t) is outdoor concentration at time *t*; *k* is the deposition rate; and *Δt* is the study sample period. *A*_*o*_ is defined as 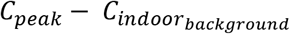.

The second model we considered was a linear regression model:

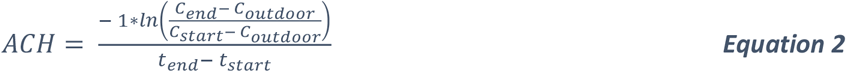

where *C*_*start*_ is the initial indoor concentration and *C*_*end*_ is the final indoor concentration; C_outdoor_ is the average outdoor concentration over the sample period; and *t*_*end*_ *− t*_*start*_ is the sample period.

The third model we considered was:

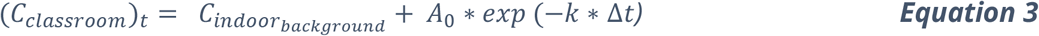

where *C*_*classroom*_ is the concentration within the classroom at time 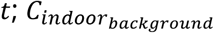 is the initial indoor concentration; *k* is the deposition rate; and *Δt* is the study sample period. *A*_*o*_ is defined as 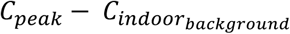.

Ultimately, we used a dynamic mass balance model (Equation 3) to calculate infiltration. This choice was based on a study on methodological approaches to estimating particle infiltration indoors. It found the dynamic model is more realistic as it accounts for particles moving in and out of an indoor microenvironment. This model assumed that there were no indoor sources, perfect mixing and no mass loss or gain due to differences in gas-phase concentrations or temperature and relative humidity conditions between indoors and outdoors.

### Indoor and outdoor concentrations

Indoor and outdoor concentrations of selected air pollutants were measured over two consecutive 24-hour time intervals following the outdoor air exchange rate measurement.

The air pollutant measurements conducted for this pilot scale study were designed to accomplish three inter-related objectives: (1) determine the outdoor air exchange rate, (2) characterize the indoor concentrations and the outdoor concentrations in the surrounding ambient air and (3) assess the effectiveness of installing a portable air cleaner in the test classroom. The instruments used to measure the pollutants of interest are presented in Table 2. The ultrafine particle instruments provide a number count concentration, not a mass concentration measurement. The black carbon devices use a light absorption method to determine the mass concentration of black carbon particles captured on an internal filter material.

**Table 2.** Air quality instruments used to measure conditions in classroom and ambient air.

| Parameter | Instrument | Manufacturer | averaging time |
| --- | --- | --- | --- |
| CO2 | LI-850 CO2 | Li-Cor Biosciences | 10 sec |
| Ultra-fine particle size distribution | NanoScan | TSI, Inc. | 1 min (full scan) |
| Particles >10 nm count | CPC | TSI, Inc. | 10 sec |
| Particles >20 nm count | P-Trak | TSI, Inc. | 10 sec |
| Black carbon | MA200 | AethLabs | 10 sec |
| Black carbon | AE51 | AethLabs | 10 sec |
| Temperature, RH | Hobo sensor | Onset Computer Corp. | 10 sec |

Classrooms were assessed twice with this research grade sampling method. At each visit, a solenoid timer valve was set to alternate 5-minute indoor and outdoor measurements with samples stored with a 10-second time resolution. Figure 3 shows the setup of these instruments for indoor/outdoor sampling.

**Figure 3.**
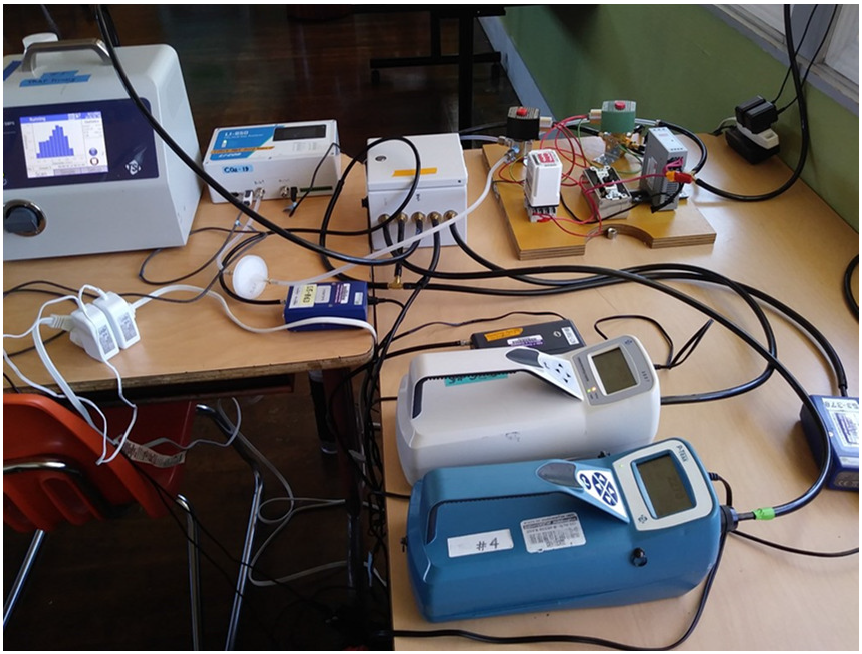
Instrument arrangement for indoor/outdoor air sampling

An inlet line to sample ambient air outside the classroom was installed using a slightly open window that was then backfilled with shim material and sealed with duct tape or by use of an available conduit to the outside from within the classroom. At the first three classroom deployments, separate instruments were used for the indoor and outdoor air sampling, but from the fourth site visit starting in June 2021, a timer and valve switch mechanism was used to alternate the inlet to the monitoring instruments between an indoor and outdoor location every 5 minutes (e.g., timer switched the valve at hh:00:00, hh:05:00, hh:10:00, etc.). The same analyzers were then measuring their respective analytes both indoors and outdoors in this alternating manner.

### Estimating infiltration by source

Each school site was visited on two occasions over the measurement period of this project. At each visit, the air quality indoors and outdoors was measured for 24 hours prior to a portable HEPA intervention and 24 hours after a HEPA filter intervention. This data provided the basis to estimate the infiltration rate of particles into the indoor space. Infiltration (see Equation 1) was calculated from 30-minute averages of indoor and outdoor concentrations and then the ratio was defined as infiltration. This required an assumption that the pollutants measured indoors were attributable to outdoor sources.

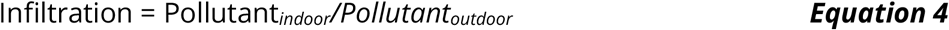

To further characterize the infiltration of pollutants by source, we used the following methods to characterize the source of the pollutants measured. Black carbon (BC) concentration was used as a proxy for diesel particle emissions, as this pollutant is primarily emitted from diesel vehicles during summer months, and our sampling did not typically overlap with wildfire events. In addition, as demonstrated in the MOV-UP study, particles with a diameter of between 10 and 20 nm are preferentially emitted by aircraft as compared to roadway traffic. Thus, the infiltration of 15.4 nm particles (measured by the NanoScan) was used to characterize the infiltration potential of aircraft emissions. Total particle number was used as a proxy for general roadway traffic. A principal component analysis (described below) was used to identify three primary features in the data, based on the correlations between the pollutants measured during each sampling period.

To estimate removal attributable to the HEPA filter unit, we calculated the ratio of infiltration after HEPA deployment as compared to the infiltration prior to HEPA deployment (Equation 2).

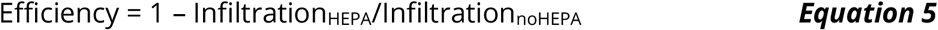

The median diameter of wildfire smoke has been shown to be centered in the 0.1 to 0.3 µm range for freshly emitted particles (with a preponderance of the mass derived from nucleation particles) and up to 1 µm for transported plumes.^59,60^

### Regression approach to removal

We also estimated the removal efficiency of the HEPA cleaner using a regression approach. A log-log multivariate linear model regressed the indoor concentration of particles to a 30-minute outdoor lagged concentration. A school-specific adjustment was used to account for differences between schools, and a term indicating the presence of HEPA filter or not was included. Based on the coefficient estimated for the HEPA term, the removal efficiency for the HEPA filter was calculated (according to Equation 2 above) across the mean observations of our study.

The log-log model output was also used to predict the concentration of particles in indoor air at the different schools when outdoor concentrations were assumed to be 5000 particles/cm^3^. Confidence intervals were generated based on propagating the error terms from the regression output.

### Principal component analysis

As previously described in the MOV-UP study^61^, a principal component analysis (PCA) was used to identify characteristic source profiles associated with the multi-pollutant data collected indoors and outdoors. Variables were created to match the input data from the MOV-UP study, and a PCA with Varimax rotation was calculated using the scaled input data. The PCA results were compared across school locations, as well as between landing and takeoff measurement conditions.

All analyses were conducted in R version 4.1.1. Packages used for analysis and output of results included data.table^62^, ggplot2^63^, emmeans^64^, zoo^65^, psych^66^, GPArotation^67^ and dplyr^68^.

## Results

The dates of sampling in the Federal Way and Highline schools are presented in Table 3, along with the flight direction of aircraft at Sea-Tac Airport relative to the school location.

**Table 3.** School classroom visit dates and aircraft operations

| School and room | First Visit | Sea-Tac flight operations overhead | Second visit | Sea-Tac flight operations |
| --- | --- | --- | --- | --- |
| School A 1<sup>st</sup> flr. | June 9-11 | Landing | July 26-28 | Takeoff |
| School A 2<sup>nd</sup> flr. | June 14-16 | Landing 14<sup>th</sup> & 15<sup>th</sup><br>Takeoff 16h | July 28-30 | Take off |
| School B | April 14-16 | Takeoff | July 20-22 | Landing 20<sup>th</sup> & 21<sup>st</sup><br>Takeoff 22<sup>nd</sup> |
| School C | April 7-9 | Takeoff | July 13-15 | Takeoff |
| School D | June 22-24 | Takeoff | August 10-12 | Landing |
| School E | March 24-26 | Takeoff 24<sup>th</sup> & 26<sup>th</sup><br>Landing 25<sup>th</sup> | July 7-9 | Takeoff |

Over the course of these deployments, 10-second data were collected both inside and outside the school using the collection of instruments described in the Methods section. This allowed for detailed information on carbon dioxide (CO_2_), black carbon (BC) and particle size to be characterized. A total of 500 MB of total data was collected over the course of the sampling period, representing 1.75 million rows of unique data. The TSI NanoScan instrument occasionally would develop operating errors over the course of the sampling. Table 4 presents a summary of the percentage of time the NanoScan instrument produced errors during the school deployments. For time periods when the NanoScan data were not available, the CPC instrument results were substituted for the total number concentration of particles (CPC does not measure multiple size ranges like the NanoScan).

**Table 4.** Percentage of missing or error flagged data

| Location | Percent Instrument Error (%) |
| --- | --- |
| School A Classroom 1, Visit 1 | 0 |
| School A Classroom 1, Visit 2 | 43 |
| School A Classroom 2, Visit 1 | 0 |
| School A Classroom 2, Visit 2 | 33 |
| School B Visit 1 | 0 |
| School B Visit 2 | 27 |
| School C Visit 1 | 0 |
| School C Visit 2 | 0 |
| School D Visit 1 | 0 |
| School D Visit 2 | 0 |
| School E Visit 1 | 19 |
| School E Visit 2 | 0 |

Outdoor exchange rates were calculated using the CO2 decay method described above. Overall, the outdoor air exchange rates ranged from 0.6/h to 4.4/h, highlighting the variability in direct exchange of air with the outdoors at the different school sites (Table 5).

**Table 5.** Outdoor air exchange rate (AER Outdoor)

| School | AER Visit 1 | AER Visit 2 |
| --- | --- | --- |
| School A |  |  |
| Room # 1 | 2.1/h | 1.3/h |
| Room # 2 | 4.4/h | 1.1/h |
| School B | 0.6/h | 0.9/h |
| School C | 2.2/h | 2.6/h |
| School D | 2.9/h | 0.4/h |
| School E | 1.1/h | 1.1/h |

### Outdoor concentration

The outdoor concentration observed at each of the five schools represents only four days of non-concurrent sampling. It is therefore difficult to directly compare the concentration of particles across the locations. Athough there were distinct differences in total pollutant concentration at the different sites, these differences are likely not representative of the year-round average differences at these sites. However, the indoor and outdoor monitoring allowed for the comparison of the infiltration dynamics over time (Figure 4).

**Figure 4.**
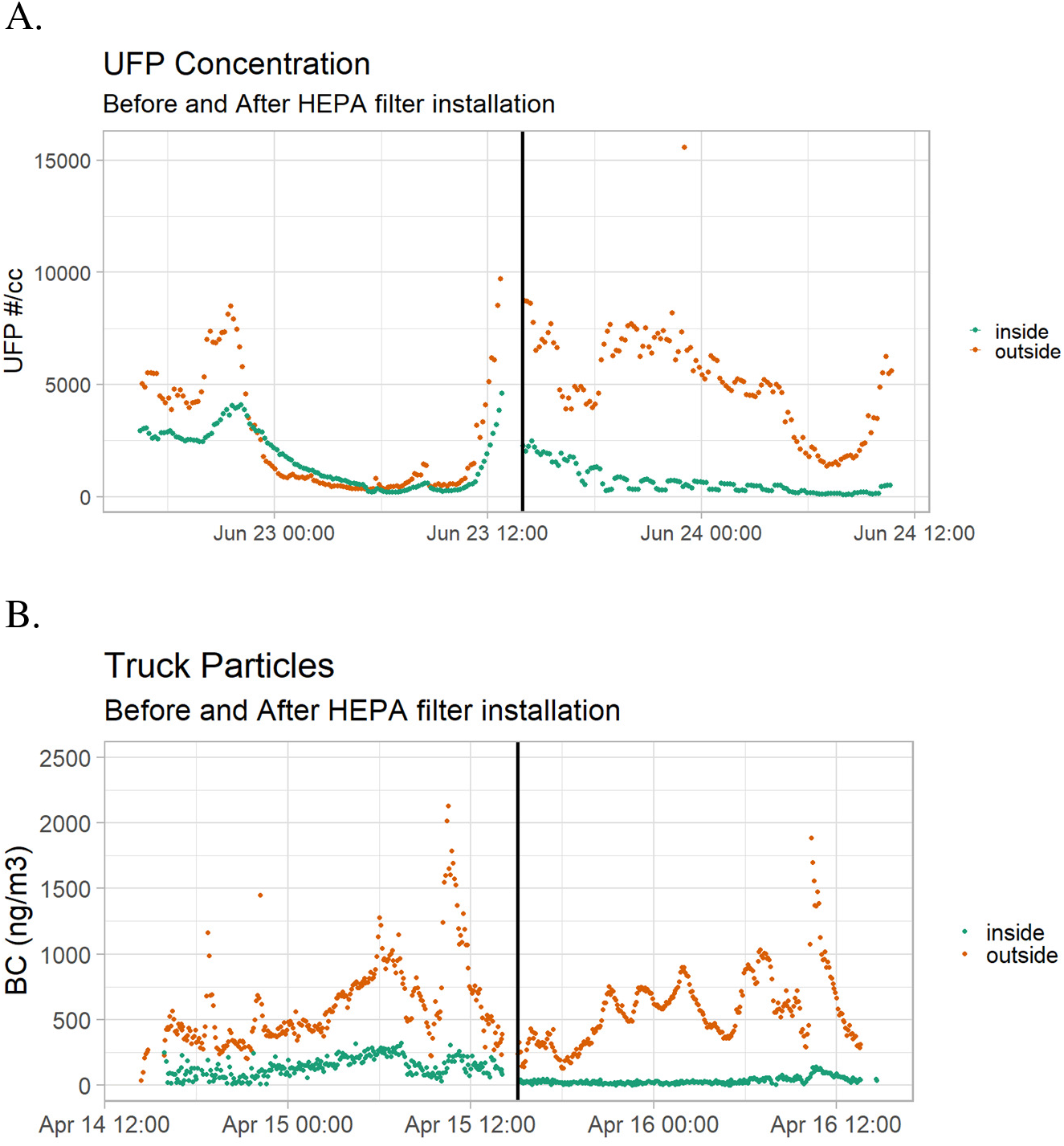
Indoor and Outdoor concentration of total particle concentration before and after portable HEPA filter deployment. B. Indoor and Outdoor concentration of Black Carbon before and after portable HEPA filter deployment.

### Observed impact of HEPA filter

The impact of the HEPA filter was observed through analysis of the relationship between the indoor and outdoor concentrations of pollutants observed over the course of deployment. As suggested in Figure 4, visual inspection suggests an effect from the use of the portable HEPA filter. Figure 6 shows an example of the change in the indoor-to-outdoor ratio of pollutants measured at the School E location, before and after the portable HEPA filter deployment. The ratio of indoor-to-outdoor air pollution was calculated for each pollutant in order to assess the impact of the HEPA filter (Figure 5).

**Figure 5.**
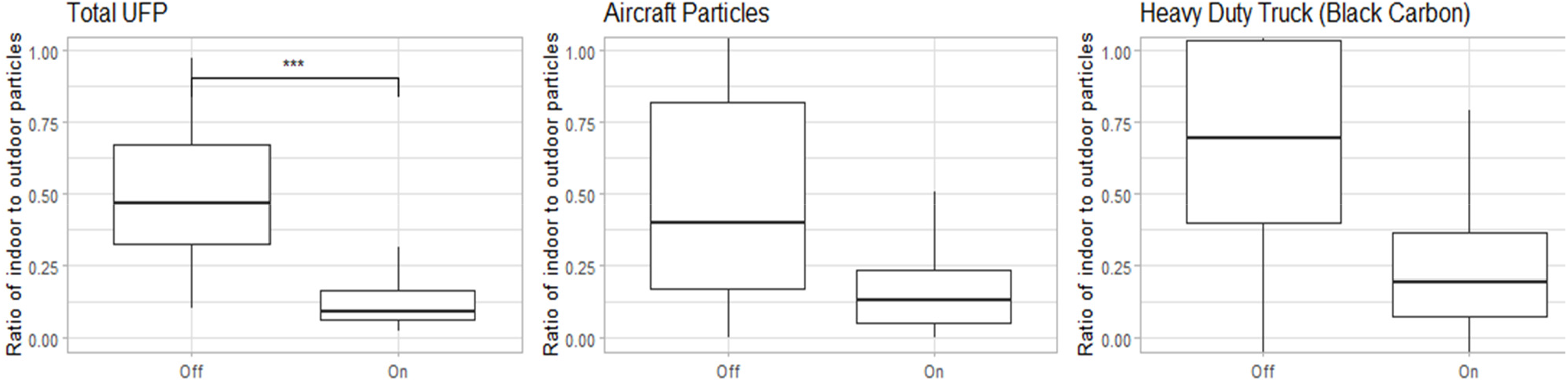
Infiltration ratio of different particle types before and after the portable HEPA filter intervention. This data represents the range of indoor/outdoor ratio of pollutants across all 5 schools. A two-sample Wilcoxon Rank Sum test confirms that the infiltration before the HEPA filter intervention is significantly higher than after the intervention (p<0.05), for each of the three particle sources.

**Figure 6.**
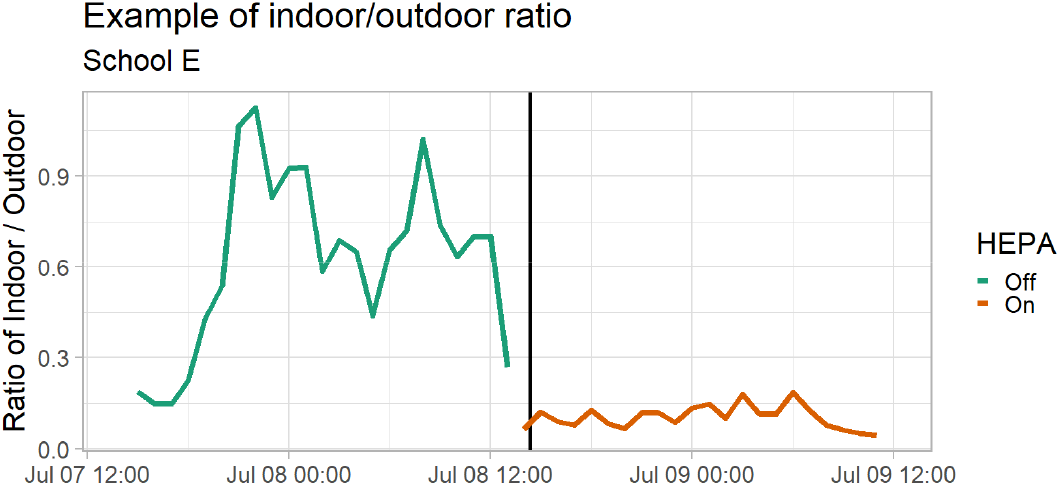
Plot if the ratio of the indoor to outdoor total particle count concentration at School E

Combining the data across all school locations, we confirm that there is a significant reduction in pollutants after the HEPA filter deployment. Table 6 presents the estimated infiltration rates with and without portable HEPA filter deployment as well as the associated confidence intervals for all estimated values. The total particle number (general traffic), particles of aircraft origin (d = 15.4 nm) and truck-traffic particles (BC) all decreased significantly after the HEPA filter deployment. Before the HEPA filter deployment, approximately half of all outdoor particles were measured indoors. After the HEPA filter deployment, approximately 1/10^th^ of all outdoor UFP were measured indoors. The removal of outdoor particles infiltrating into the indoor space attributed to the portable HEPA filter is estimated to be 83% removal for UFP, 67% removal for aircraft particles and 73% removal for heavy-duty traffic particles. This represents a removal efficiency of 83% for removal of particles of outdoor origin (Equation 5). The estimated median removal indoors is not significantly different among particle types, suggesting that the HEPA filter intervention is effective for all outdoor air pollutants, including aircraft, wildfire and roadway particles.

**Table 6.** Infiltration (%) with and without the portable HEPA filter unit installed in classrooms

| **Pollutant Type** | **Infiltration before HEPA** | **Confidence Range (%)** | **Infiltration After HEPA** | **Confidence Range** | **Removal by HEPA (%)** | **Confidence Range** |
| --- | --- | --- | --- | --- | --- | --- |
| **Total UFP** | 54% | [47, 59] | 9% | [8, 9] | 83% | [82,84] |
| **Aircraft Particles** | 41% | [38, 56] | 14% | [12, 15] | 67% | [68, 73] |
| **Heavy Duty Truck** | 74% | [71, 79] | 20% | [18,21] | 73% | [73, 74] |

We also calculated Pearson’s Correlation Coefficient to better understand the relationship between indoor and outdoor concentrations with and without the HEPA filter deployed. We found that without the HEPA filter, there was a 40% correlation (moderate) between the indoor and outdoor measures. When the HEPA filter was deployed, there was a 9% correlation (weak) between the indoor and outdoor measures. We also found that this relationship between indoor and outdoor air quality persisted for lags of up to 60 minutes without a HEPA filter, but that there was no observable correlation between indoor and outdoor when the HEPA filter was introduced. This can be observed in Figure 4 where the indoor concentration closely follows the change in outdoor concentration before the introduction of the HEPA filter (moderate correlation). After the introduction of the HEPA filter, there is no obvious relationship between the change in outdoor concentration and the change in indoor concentration (poor correlation).

### Modeled impact of the HEPA filter

In order to better understand the overall impact of HEPA filtration, we developed a model to predict indoor concentration based on the school location, use of a HEPA filter and average outdoor concentration over the previous 30 minutes. This model assumed that the indoor concentration represented a fraction of the outdoor concentration (log-log model). We then predicted the average indoor air quality concentration at each school, for a fixed outdoor concentration of 5,000 #/cm^3^ with and without the HEPA filter intervention. We saw a statistically significant decrease in indoor air quality concentration in all the schools, with School A having the highest infiltration rates with and without the HEPA filter intervention (Figure 7). School E was not included in the model as there were multiple indoor concentrations values of zero observed after the HEPA filter deployment, making it impossible to include this location in the log-log model. Overall, we estimated that the HEPA filter effectiveness was 71% [70%-72%] across the measurement conditions, after accounting for school-specific differences. This regression result is consistent with the result observed when calculating HEPA filter effectiveness using the ratio of indoor-to-outdoor pollutants (Table 6). In Phase 2, this model will be further expanded to include information on building age and ventilation type.

**Figure 7.**
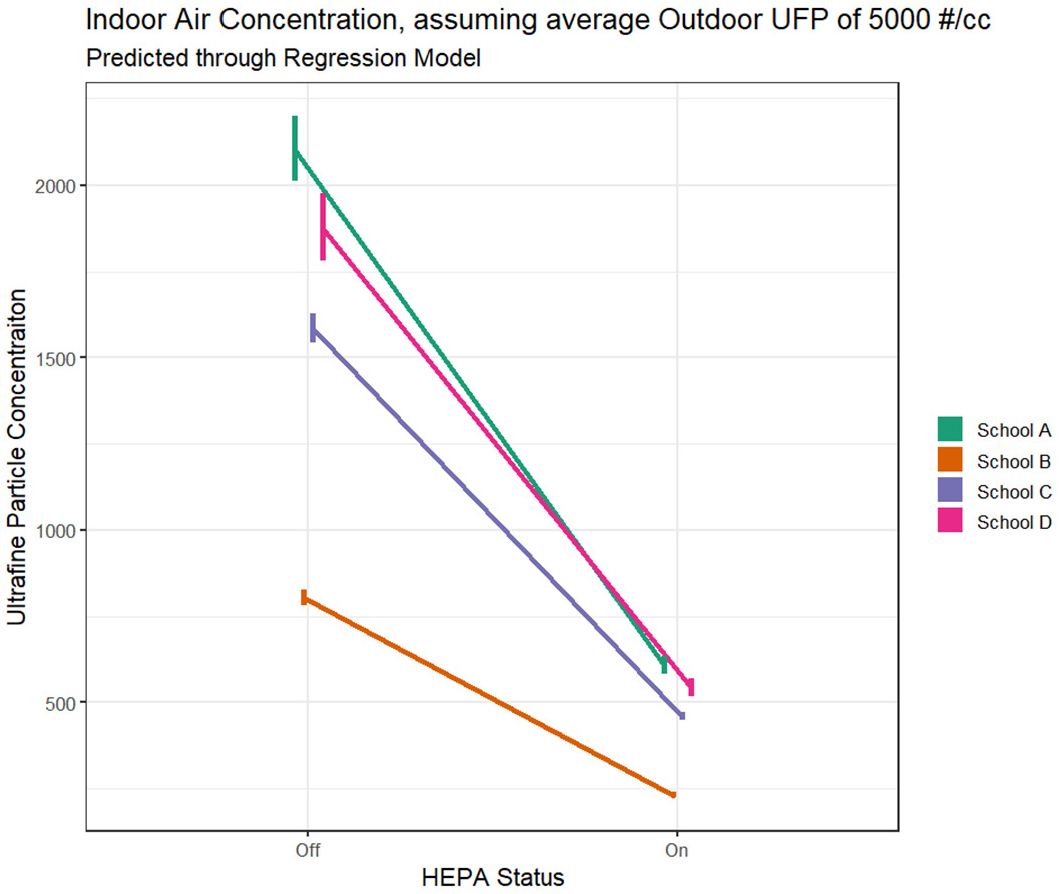
Prediction of indoor concentrations with and without a portable HEPA filter deployed in the classroom. School E was not included in this model due to multiple 0 values for the indoor air quality measurement.

**Figure 8.**
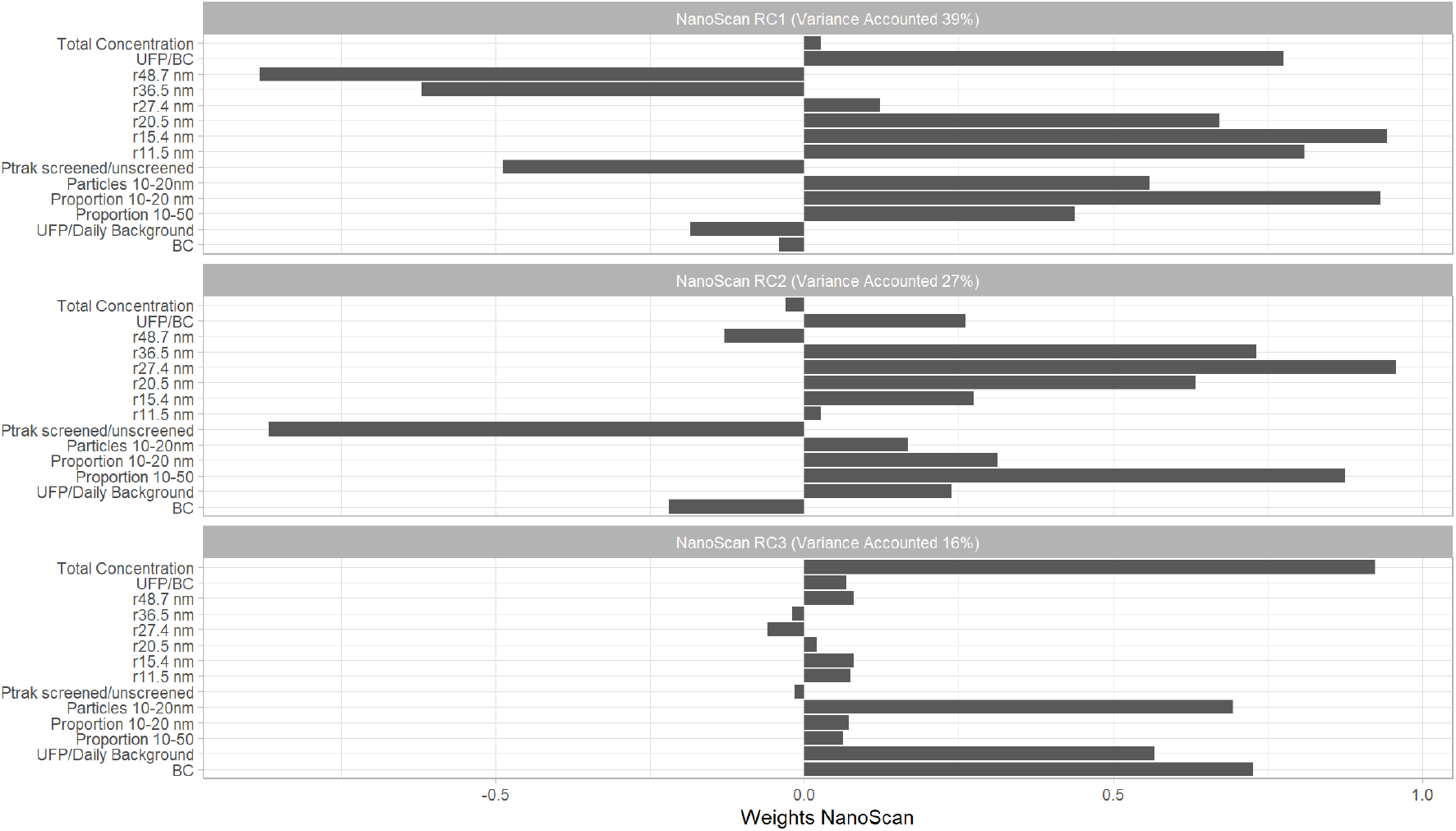
PCA score loadings after Varimax rotation. These 3 PCA components account for 82% of the variability observed in the outdoor air pollution data collected. The source of each component is based on the loadings onto each variable that entered in the model. The loadings observed in this analysis, were very similar to the loadings observed in MOV-UP allowing for the components to be labeled as: RC1 – Aircraft, RC2 – General Roadway and RC3 – Heavy duty truck.

### Principal Component Analysis

We selected three components for the Principal Component Analysis (PCA). As in the previous MOV-UP study, the components were selected by examination of a scree plot. The 3 components selected were analogous to the three components generated in the MOV-UP study when using NanoScan data. These components accounted for 82% of the observed variability. The first component was consistent with the aircraft feature identified in MOV-UP, the second component consistent with the roadway traffic feature and the third consisted with the high diesel feature.

We compared the distribution of component scores outdoors at the different school locations (Figure 10). We noted significant variability in the strength of the components, with roadway contributing most significantly to the observed pollutant matrix, followed by aircraft and truck diesel. Although some differences were observed in the distribution within PCA component at each school, there were no significant differences noted in this data, except for School B, which had a significantly higher contribution of truck diesel.

We also used the results of the PCA to examine the impact of aircraft landing overhead on the contribution of our PCA features to the total outdoor concentration of particles (Figure 9). In order to capture periods of time when there were multiple source contributions to the ultrafine particle concentration, we restricted this analysis to data points when the total concentration of particles was greater than 5,000 #/cm^3^. We then plotted the scores of the PCA components for takeoff and landing conditions (see Table 3). The results presented in Figure 9 show that only the aircraft component is significantly increased during periods of aircraft landing overhead. The other components do not show a statistically significant change in contribution to the total particle concentration.

**Figure 9.**
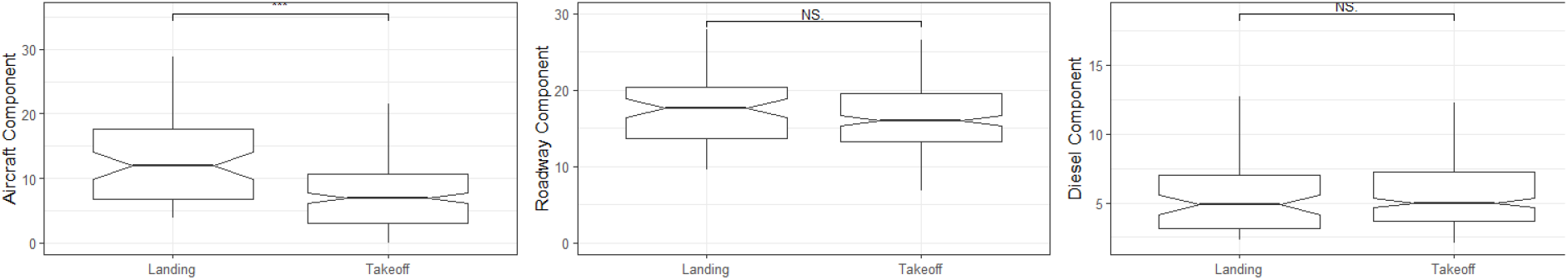
Contribution of each source feature to the outdoor total particle number concentration during Landing and Takeoff overhead conditions. The contribution of aircraft sources is significantly higher in Landing conditions. The aircraft landing direction is not significantly associated with change in the roadway and diesel features.

**Figure 10.**
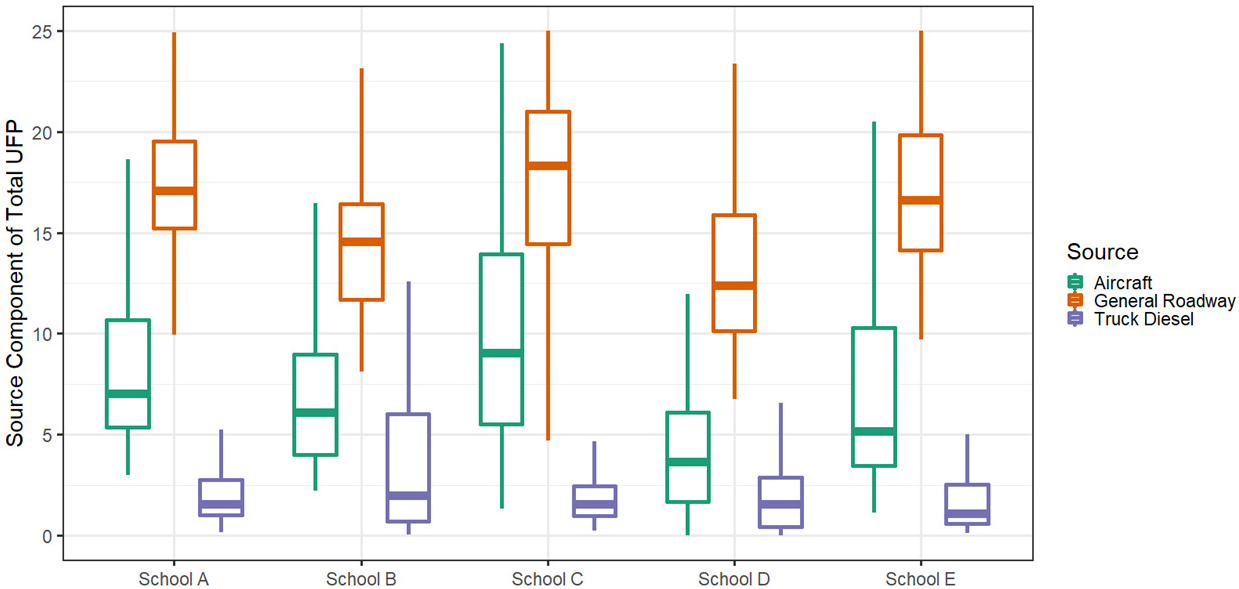
PCA component score distribution over the course of sampling at the 3 schools.

This result suggests that aircraft landing overhead do contribute a significant proportion of 10-20 nm particles to the overall composition of particles measured at outdoor locations, particularly in proximity to flight paths. All of the schools selected for this project were within 0.5 miles of a Sea-Tac flight path. Additionally, we find that there are significant impacts of general roadway traffic as well as specific heavy-duty diesel traffic at all the school locations sampled.

In order to confirm that the particles measured were primarily of outdoor origin, we predicted the PCA component scores for the indoor data collected, based on the model developed from the outdoor-only data. We then looked at the distribution in PCA scores across schools, both indoor and outdoor (Figure 11). We observe that for all school locations, the distribution of scores is fairly consistent between the indoor and outdoor data. For the diesel component, there is evidence that there are significantly lower scores indoors. For the other PCA scores, there is no systematic significant difference between the indoor and outdoor scores. This suggests that the particles measured indoors are attributable to both aircraft and roadway traffic emissions and are unlikely to be generated indoors by other sources.

**Figure 11.**
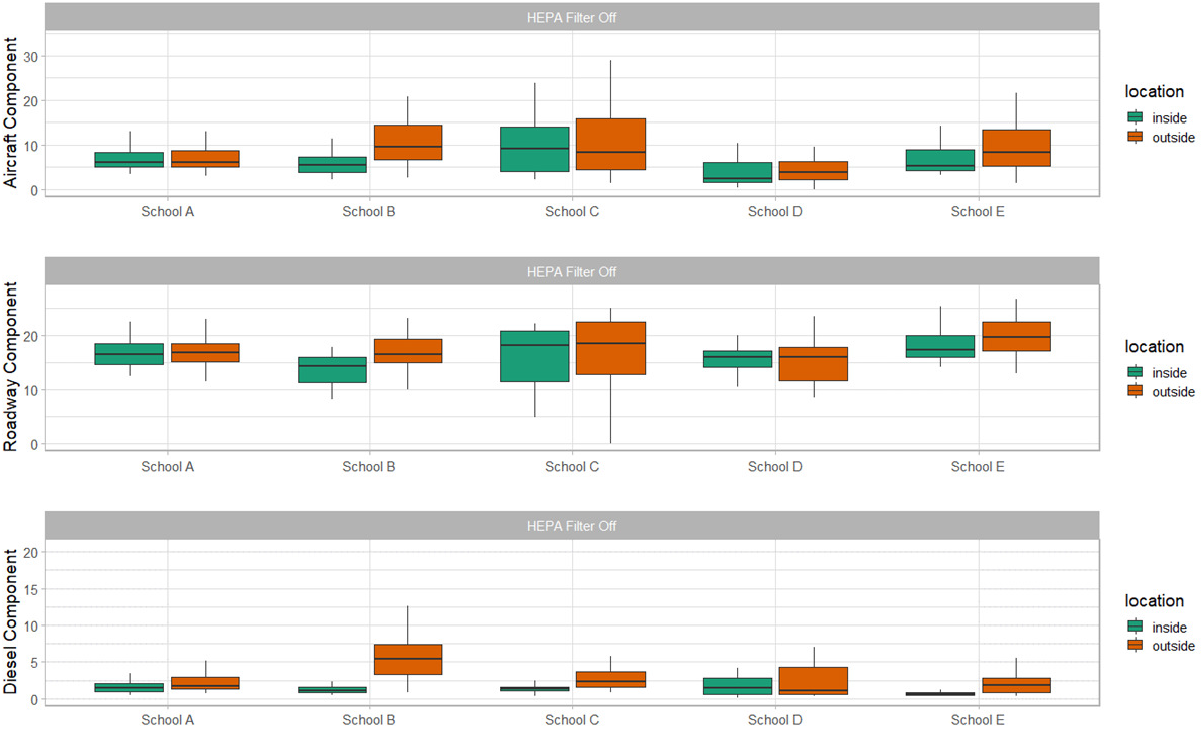
PCA contribution indoor and outdoor at the 5 school locations. There are significant decreases in indoor contribution of diesel traffic at all the schools. For the aircraft and roadway components, there is no significant reduction in the scores observed indoors.

### Overall distribution of pollutants

We find that prior to HEPA filter deployment, concentrations of ultrafine particles, ultra-ultrafine particles and black carbon outdoors are significantly higher than those measured indoors. This is consistent for all measured pollutants. We consistently find that the total concentrations measured indoors are lower after the HEPA filter deployment, as shown below. Consistent with the findings of this paper that the portable HEPA filter has a significant impact on indoor air quality, we observe a reduction of the pollution from outdoor sources persisting in the classroom environment (Figure 12). The indoor/outdoor ratio also varied by school location. In Figure 13 we present the results of the observed 30-minute indoor/outdoor ratios at each school location. Consistently, there are lower ratios after the HEPA filter deployment, but the magnitude of this change varies by school.

**Figure 12.**
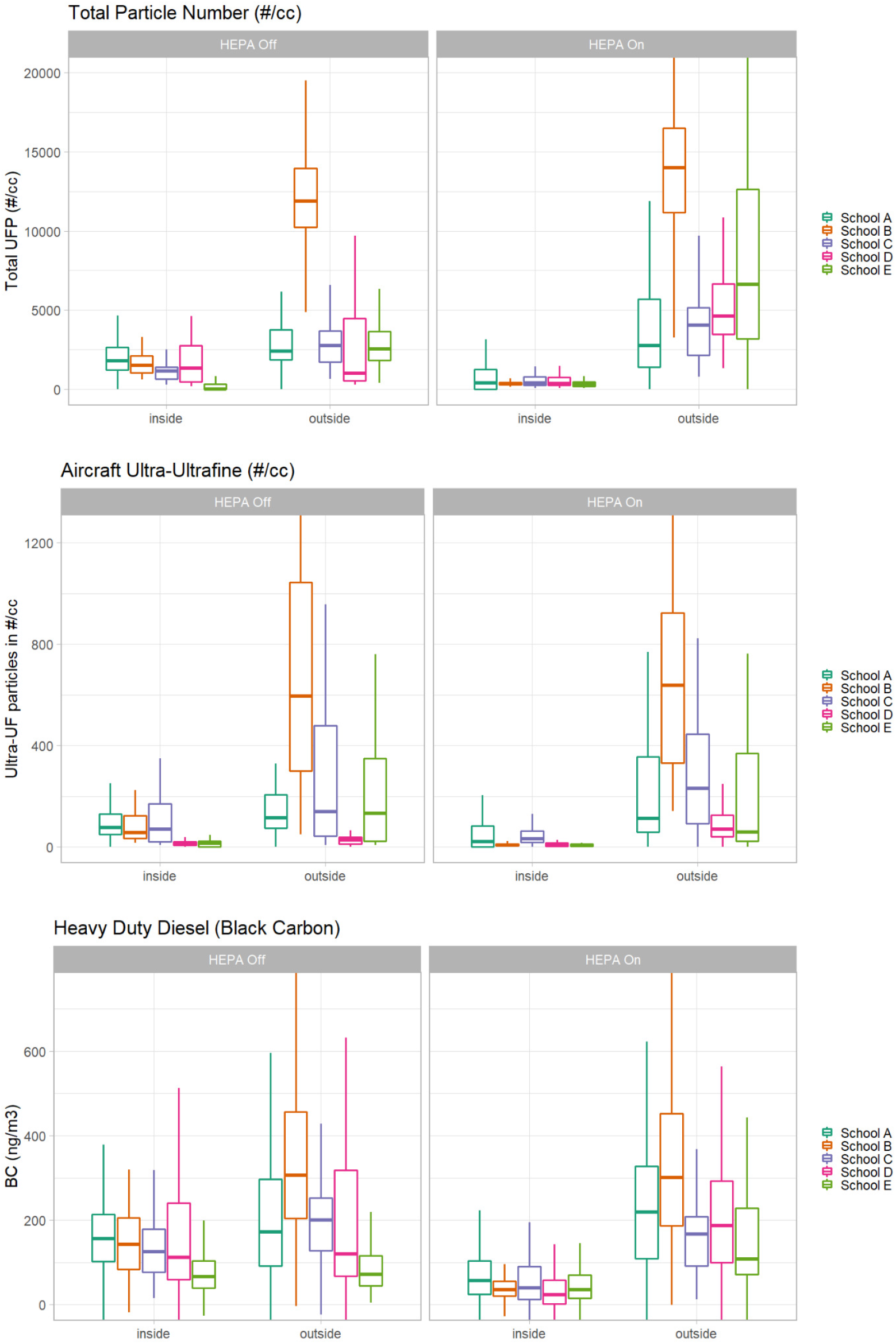
Distribution of Pollutants before and after HEPA filter deployment at all 5 school locations

**Figure 13.**
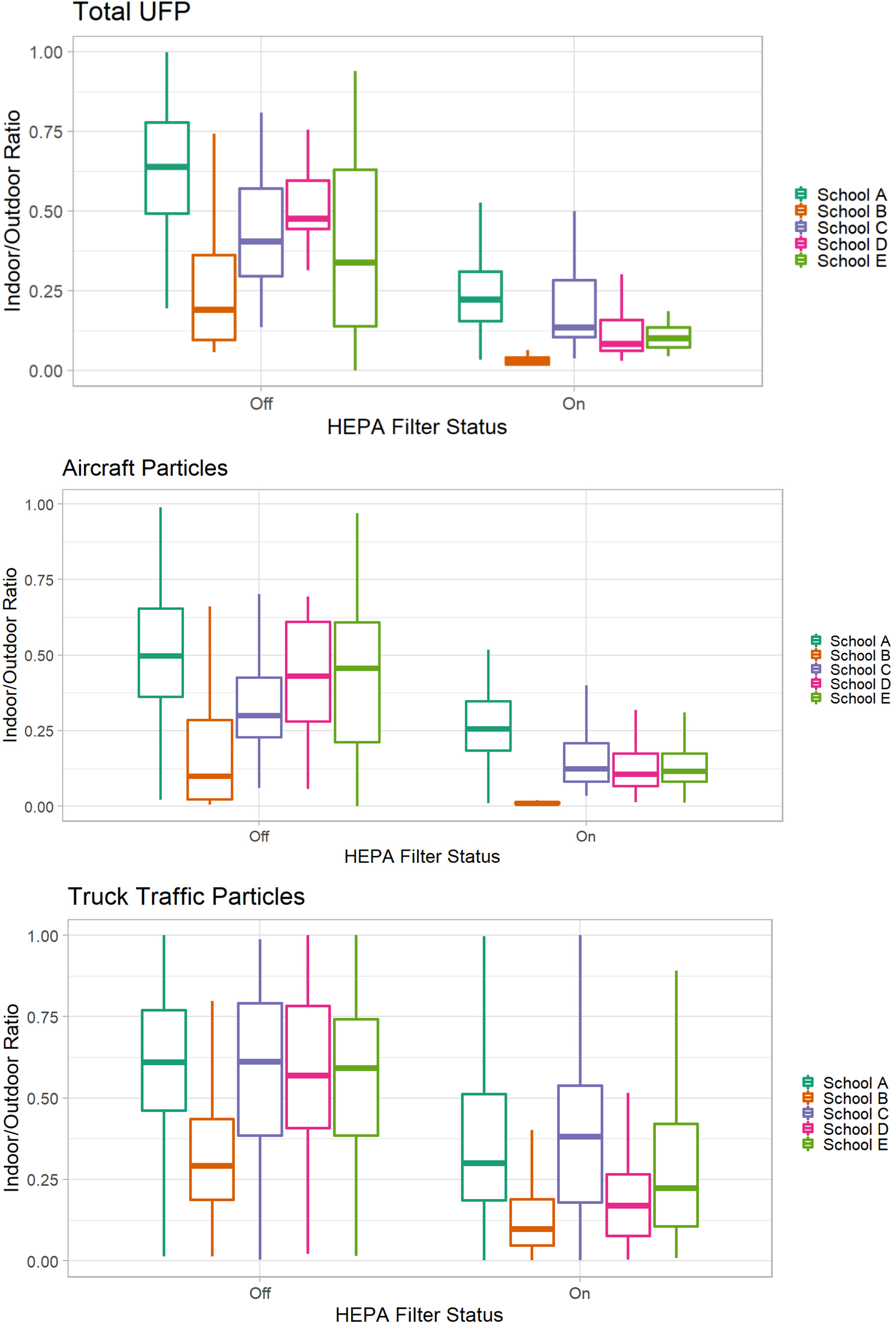
Distribution indoor/outdoor ratio values at each school locations. Values were compared on the 30-minute timescale.

## Discussion

Ultrafine particles are not routinely measured in the outdoor environment by air quality agencies across the United States. However in recent years, there have been special studies in Pittsburgh, the Netherlands, New York, Montreal, Seattle and Los Angeles that confirm UFPs are elevated near roadways, near industrial sites, in urban cores and in proximity to flight paths.^2,69–71^ There are strong gradients of exposure to UFPs observed in these studies, with UFP decreasing to background levels within 100 meters of sources. Because of the lack of health-based regulatory standards as well as limited long-term monitoring data, it is difficult to compare the magnitude of the outdoor concentrations observed in this study to typical outdoor concentrations.

To inventory available regulatory UFP monitoring data, we searched the EPA Air Quality System (AQS) database and contacted select local air quality agencies across the US. We found some form of UFP monitoring data near Baltimore, Miami, New York, Saint Paul, Pittsburgh, Los Angeles and Seattle. In general, these special studies were either designed as short-term mobile monitoring studies or snapshot designs, where monitors are rotated among fixed sites for a year or less.^2,72–75^ The New York State Department of Environmental Conservation (DEC) collected 1-minute UFP count data across seven sites in New York State over the year 2017 at near-road, urban, suburban and state park locations. This dataset was shared with the research team. The dataset clearly demonstrates UFP gradients away from roadway, with a site located directly next to a freeway in Queens, NY, reporting 1.5 to 2 times greater concentrations of UFP than a site located only 300 m downwind of the road. When comparing the data collected in this study to the data from NY, the concentrations of UFP measured at School B was comparable to the concentration of UFP measured at near-roadway sites in Buffalo and Rochester, NY (median concentration at School B was 12,050 #/cc, and the median concentration at the near-roadway sites in NY was 14,000 #/cc). The median concentration at the four other school locations were lower and more similar to urban concentration observed at sites not located directly next to major roadways in NY. Because the measurements in this study were only conducted over a total of four days, it would be informative to continue longer-term monitoring to better understand and quantify the impact of UFP across this area.

Infiltration rates observed in this project are approximately 50%, meaning about half of outdor pollution is entering into classroom spaces without any additional portable HEPA filtration. A study in Boston, MA, looked at the infiltration of aircraft-related UFP into residential buildings in proximity to flight paths.^76^ Hudda et al. found that median outdoor concentrations of UFP were 19,000 #/cc when wind direction placed the residence downwind of the flight path and 10,000 #/cc during other wind conditions. The authors also found significant infiltration of aircraft particles into local residences and calculated a 33% decrease in indoor concentration after a portable HEPA filter was installed. These findings are consistent with the findings in this study, although HEPA filter effectiveness and infiltration rates were not calculated in that project.

Recent controlled interventions have established improvements in symptoms of children with asthma after a HEPA filter intervention in their homes. ^77–80^ These studies also show consistent improvement in the indoor air quality of these homes after HEPA filter intervention. However, none of these studies directly evaluated UFP, the primary pollutant of interest in this study, instead focusing on the PM_2.5_ fraction of air pollution.

Studies evaluating the impact of HEPA filtration in school settings are limited. A recently conducted randomized trial of asthmatic children receiving a HEPA filter intervention combined with integrated pest management concluded that there were significant improvements to indoor air quality, with a 45% reduction in indoor PM_2.5_ in HEPA-treated classrooms as compared to untreated classrooms.^81^ However, this study did not find improvement in asthma symptoms for the 118 asthmatic students receiving HEPA filtration as compared to the 118 students not receiving HEPA filtration. These 236 students were attending 41 different schools. Several limitations were highlighted by the authors, including a lack of fine-scale data on exposure at each school during the study period as well as masking of an effect by seasonal viral illnesses.

A randomized crossover study of HEPA filtration, without a washout period, in 23 homes of low-income Puerto Ricans in Boston and Chelsea, MA, concluded that a portable HEPA filter intervention resulted in significant improvement of indoor air quality.^82^ Median UFP when using HEPA filtration was 50% to 85% lower compared to nonfiltration in most homes.

Although this study also measured health outcomes, there was no observed benefit to HEPA filtration in terms of reduced inflammation. In the Healthy Air, Healthy Schools Phase 1 project, we estimated that HEPA filtration resulted in 70% to 80% lower UFP as compared to no additional filtration. This result is consistent with the findings from Boston. Although the Boston-based pilot project did not observe inflammation reduction in their study population, possible limitations of the study include a study population that is unresponsive to UFP due to general good health as well as not enough enrolled participants to observe an effect.

The findings of the Healthy Air, Healthy Schools Project Phase 1 are consistent with existing literature demonstrating that HEPA filter interventions reduce exposure to outdoor pollutants in indoor spaces. This study is unique in focusing on UFP in school settings and demonstrating through multivariate methods that the UFP measured in the classroom space is primarily of outdoor origin. Although existing research suggests that improvements to indoor air quality in homes can significantly improve asthma outcomes, further research is necessary to establish the benefit to student health and academic performance of improved air quality in schools.

## Conclusions and recommendations

Indoor air quality in schools is significantly impacted by outdoor sources of ultrafine particles. Portable HEPA filters can significantly reduce the concentration of outdoor pollution in the classroom. Using portable HEPA filter units reduced indoor concentrations of UFP by approximately 70%. The main findings and recommendations that emerge from Phase 1 of this project are:

1. Schools that are near truck routes, aircraft flight paths and high-traffic roadways are at higher risk of indoor air pollution.
2. Landing aircraft contribute significantly to indoor and outdoor UFP concentrations in this study region.
3. Portable HEPA filter units can be effectively used in the short term to decrease air pollution in a classroom space by removing particles.
4. Ventilation changes and building-level remediations such as sealing gaps and managing doorways should be investigated as an approach to reduce infiltration of outdoor particles indoors.
5. The optimal usage of HEPA filter units will be evaluated in Phase 2 of this project to balance energy usage and air quality management.
6. The health and well-being benefits of reducing UFP concentrations indoors must still be investigated in Phase 2.
7. The methodology to identify schools at higher risk of UFP impacts will be developed in Phase 2.

## Data Availability

All data produced in the present study are available upon reasonable request to the authors.

## References

1. Johnson, K., Solet, D. & Serry, K. Community Health and Airport Operations Related Noise and Air Pollution; Report to the Legislature in Response to Washington State HOUSE BILL 1109. (Public Health Seattle & King County; Assessment, Policy Development and Evaluation Unit, 2020).

2. Austin, E. et al. Distinct Ultrafine Particle Profiles Associated with Aircraft and Roadway Traffic. Environ. Sci. Technol. 55, 2847–2858 (2021).

3. Blanco, M. N. Traffic-Related Air Pollution and Dementia Incidence in a Seattle-Based, Prospective Cohort Study. (2021).

4. Blanco, M. N. et al. Mobile monitoring of traffic-related air pollution for a prospective cohort study in the greater Seattle area. medRxiv (2021) doi:https://doi.org/10.1101/2021.09.18.21263522.

5. Westerdahl, D., Fruin, S., Sax, T., Fine, P. M. & Sioutas, C. Mobile platform measurements of ultrafine particles and associated pollutant concentrations on freeways and residential streets in Los Angeles. Atmos Env. 39, 3597–3610 (2005).

6. Hudda, N., Simon, M., Zamore, W. & Durant, J. Aviation-Related impacts on ultrafine particle number concentrations outside and inside residences near an airport. Env. Sci Technol 52, 1765–1772 (2018).

7. Shirmohammadi, F. et al. Emission rates of particle number, mass and black carbon by the Los Angeles International Airport (LAX) and its impact on air quality in Los Angeles. Atmos Env. 151, 82–93 (2017).

8. Hudda, N. & Fruin, S. International airport impacts to air quality: size and related properties of large increases in ultrafine particle number concentrations. Env. Sci Technol 50, 3362– 3370 (2016).

9. Riley, E. A. et al. Ultrafine particle size as a tracer for aircraft turbine emissions. Atmos Env. 139, 20–29 (2016).

10. Hudda, N., Simon, M., Zamore, W., Brugge, D. & Durant, J. Aviation emissions impact ambient ultrafine particle concentrations in the greater Boston area. Env. Sci Technol 50, 8514–8521 (2016).

11. Masiol, M. et al. Analysis of major air pollutants and submicron particles in New York City and Long Island. Atmos Env. 148, 203–214 (2017).

12. Keuken, M. P., Moerman, M., Zandveld, P., Henzing, J. S. & Hoek, G. Total and size-resolved particle number and black carbon concentrations in urban areas near Schiphol airport (the Netherlands). Atmos. Environ. 104, 132–142 (2015).

13. Larson, T. et al. Ambient air quality measurements from a continuously moving mobile platform: Estimation of area-wide, fuel-based, mobile source emission factors using absolute principal component scores. Atmos Env. 152, 201–211 (2017).

14. Keller, J. P. et al. Pollutant composition modification of the effect of air pollution on progression of coronary artery calcium: the Multi-Ethnic Study of Atherosclerosis. Environ. Epidemiol. Phila. Pa 2, (2018).

15. Wing, S. E. et al. Preterm Birth among Infants Exposed to in Utero Ultrafine Particles from Aircraft Emissions. Environ. Health Perspect. 128, 047002 (2020).

16. Qian, Z. et al. Ambient air pollution and preterm birth: A prospective birth cohort study in Wuhan, China. Int. J. Hyg. Environ. Health 219, 195–203 (2016).

17. Habre, R. et al. Short-term effects of airport-associated ultrafine particle exposure on lung function and inflammation in adults with asthma. Environ. Int. 118, 48–59 (2018).

18. Holm, S. M., Miller, M. D. & Balmes, J. R. Health effects of wildfire smoke in children and public health tools: a narrative review. J. Expo. Sci. Environ. Epidemiol. (2020) doi:10.1038/s41370-020-00267-4.

19. Rim, D., Wallace, L. & Persily, A. Infiltration of Outdoor Ultrafine Particles into a Test House. Environ. Sci. Technol. 44, 5908–5913 (2010).

20. Long, C. M., Suh, H. H., Catalano, P. J. & Koutrakis, P. Using Time-and Size-Resolved Particulate Data To Quantify Indoor Penetration and Deposition Behavior. Environ. Sci. Technol. 35, 2089–2099 (2001).

21. Zweig, J. S., Ham, J. C. & Avol, E. L. Air pollution and academic performance: Evidence from California schools. (National Institute of Environmental Health Sciences, 2009).

22. Stafford, T. M. Indoor air quality and academic performance. J. Environ. Econ. Manag. 70, 34–50 (2015).

23. Gilraine, M. Air Filters, Pollution and Student Achievement. (2020) doi:10.26300/7MCR-8A10.

24. Dockery, D. W. & Pope, C. A. Acute respiratory effects of particulate air pollution. Annu. Rev. Public Health 15, 107–132 (1994).

25. United States Environmental Protection Agency. Integrated Science Assessment (ISA) for Particulate Matter. https://cfpub.epa.gov/ncea/risk/recordisplay.cfm?deid=216546 (2009).

26. United States Environmental Protection Agency. Criteria Air Pollutants (America’s Children and the Environment). (2015).

27. Calderón-Garcidueñas, L. et al. Air pollution and brain damage. Toxicol. Pathol. 30, 373– 389 (2002).

28. Calderón-Garcidueñas, L. et al. Long-term air pollution exposure is associated with neuroinflammation, an altered innate immune response, disruption of the blood-brain barrier, ultrafine particulate deposition, and accumulation of amyloid beta-42 and alpha-synuclein in children and young adults. Toxicol. Pathol. 36, 289–310 (2008).

29. Calderón-Garcidueñas, L. et al. Neuroinflammation, hyperphosphorylated tau, diffuse amyloid plaques, and down-regulation of the cellular prion protein in air pollution exposed children and young adults. J. Alzheimers Dis. JAD 28, 93–107 (2012).

30. Tessum, C. W. et al. Inequity in consumption of goods and services adds to racial–ethnic disparities in air pollution exposure. Proc. Natl. Acad. Sci. 116, 6001–6006 (2019).

31. Adams, C. et al. Disentangling the Exposure Experience: The Roles of Community Context and Report-back of Environmental Exposure Data. J. Health Soc. Behav. 52, 180–196 (2011).

32. Claudio, L., Gilmore, J., Roy, M. & Brenner, B. Communicating environmental exposure results and health information in a community-based participatory research study. BMC Public Health 18, 784 (2018).

33. Pope, C. A. & Dockery, D. W. Health effects of fine particulate air pollution: lines that connect. J. Air Waste Manag. Assoc. 1995 56, 709–742 (2006).

34. United States Environmental Protection Agency. Integrated Science Assessment (ISA) for Particulate Matter. (2019).

35. Power, M. C., Adar, S. D., Yanosky, J. D. & Weuve, J. Exposure to air pollution as a potential contributor to cognitive function, cognitive decline, brain imaging, and dementia: A systematic review of epidemiologic research. Neurotoxicology 56, 235–253 (2016).

36. Tzivian, L. et al. Long-term air pollution and traffic noise exposures and cognitive function:A cross-sectional analysis of the Heinz Nixdorf Recall study. J. Toxicol. Environ. Health A 79, 1057–1069 (2016).

37. Crouse, D. L. et al. Risk of nonaccidental and cardiovascular mortality in relation to long-term exposure to low concentrations of fine particulate matter: a Canadian national-level cohort study. Environ. Health Perspect. 120, 708–714 (2012).

38. Pinault, L. et al. Risk estimates of mortality attributed to low concentrations of ambient fine particulate matter in the Canadian community health survey cohort. Environ. Health Glob. Access Sci. Source 15, 18 (2016).

39. Government of Canada. The Georgia Basin-Puget Sound Airshed Characterization Report 2014: chapter 5. https://www.canada.ca/en/environment-climate-change/services/air-pollution/publications/georgia-basin-puget-sound-report-2014/chapter-5.html (2014).

40. Washington Department of Health. Outdoor Air Quality. https://www.doh.wa.gov/CommunityandEnvironment/AirQuality/OutdoorAir#FineParticulateMatter (2018).

41. Puget Sound Clean Air Agency. 2019 Air Quality Data Summary. (2020).

42. City of Seattle. Air Quality and Greenhouse Gas Emissions.

43. Puget Sound Clean Air Agency. Criteria Air Pollutants. https://www.pscleanair.org/163/Criteria-Air-Pollutants (2018).

44. Halofsky, J. E., Peterson, D. L. & Harvey, B. J. Changing wildfire, changing forests: the effects of climate change on fire regimes and vegetation in the Pacific Northwest, USA. Fire Ecol. 16, 4 (2020).

45. Goss, M. et al. Climate change is increasing the likelihood of extreme autumn wildfire conditions across California. Environ. Res. Lett. 15, 094016 (2020).

46. Doubleday, A. et al. Mortality associated with wildfire smoke exposure in Washington state, 2006–2017: a case-crossover study. Environ. Health 19, 4 (2020).

47. Washington Department of Health. Smoke From Fires. https://www.doh.wa.gov/CommunityandEnvironment/AirQuality/SmokeFromFires#headingp49513 (2021).

48. Lammers, A. et al. Effects of short-term exposures to ultrafine particles near an airport in healthy subjects. Environ. Int. 141, 105779 (2020).

49. Hudda, N., Simon, M. C., Zamore, W., Brugge, D. & Durant, J. L. Aviation Emissions Impact Ambient Ultrafine Particle Concentrations in the Greater Boston Area. Environ. Sci. Technol. 50, 8514–8521 (2016).

50. Klepeis, N. E. et al. The National Human Activity Pattern Survey (NHAPS): a resource for assessing exposure to environmental pollutants. J. Expo. Anal. Environ. Epidemiol. 11, 231– 252 (2001).

51. United States Environmental Protection Agency, O. Why Indoor Air Quality is Important to Schools. https://www.epa.gov/iaq-schools/why-indoor-air-quality-important-schools (2015).

52. Hemond, H. F. & Fechner, E. J. Chapter 4 - The Atmosphere. in Chemical Fate and Transport in the Environment (Third Edition) (eds. Hemond, H.F. & Fechner, E.J.) 311– 454 (Academic Press, 2015). doi:10.1016/B978-0-12-398256-8.00004-9.

53. Cavaleiro Rufo, J. et al. Children exposure to indoor ultrafine particles in urban and rural school environments. Environ. Sci. Pollut. Res. 23, 13877–13885 (2016).

54. García-Hernández, C., Ferrero, A., Estarlich, M. & Ballester, F. Exposure to ultrafine particles in children until 18 years of age: A systematic review. Indoor Air 30, 7–23 (2020).

55. Chen, Y. & Chen, B. The Combined Effect of Indoor Air Quality and Socioeconomic Factors on Health in Northeast China. Appl. Sci. 10, 2827 (2020).

56. United States Environmental Protection Agency, O. What is a HEPA filter? https://www.epa.gov/indoor-air-quality-iaq/what-hepa-filter-1 (2019).

57. Azimi, P., Zhao, D. & Stephens, B. Estimates of HVAC filtration efficiency for fine and ultrafine particles of outdoor origin. Atmos. Environ. 98, 337–346 (2014).

58. Allen, J., Spengler, J., Jones, E. & Cedeno-Laurent, J. 5-step guide to checking ventilation rates in classrooms. (Harvard Healthy Buildings Program, 2020).

59. Janhäll, S., Andreae, M. O. & Pöschl, U. Biomass burning aerosol emissions from vegetation fires: particle number and mass emission factors and size distributions. Atmospheric Chem. Phys. 10, 1427–1439 (2010).

60. Radke, L. F. et al. Airborne monitoring and smoke characterization of prescribed fires on forest lands in western Washington and Oregon. PNW-GTR-251 https://www.fs.usda.gov/treesearch/pubs/5623 (1990) xdoi:10.2737/PNW-GTR-251.

61. Austin, E. et al. Mobile ObserVations of Ultrafine Particles: The MOV-UP study report. (2019).

62. Dowle, M. et al. data.table: Extension of ‘data.frame’. (2021).

63. Wickham, H. ggplot2: ggplot2. Wiley Interdiscip. Rev. Comput. Stat. 3, 180–185 (2011).

64. Searle, S. R., Speed, F. M. & Milliken, G. A. Population Marginal Means in the Linear Model: An Alternative to Least Squares Means. Am. Stat. 34, 216–221 (1980).

65. Zeileis, A. & Grothendieck, G. zoo: S3 Infrastructure for Regular and Irregular Time Series. arXiv:math/0505527 (2005).

66. Revelle, W. psych: Procedures for Psychological, Psychometric, and Personality Research. (2021).

67. Jennrich, C. B. and R. GPArotation: GPA Factor Rotation. (2014).

68. Wickham, H., François, R., Henry, L., Müller, K. & RStudio. dplyr: A Grammar of Data Manipulation. (2021).

69. Saha, P. K., Hankey, S., Marshall, J. D., Robinson, A. L. & Presto, A. A. High-Spatial-Resolution Estimates of Ultrafine Particle Concentrations across the Continental United States. Environ. Sci. Technol. acs.est.1c03237 (2021) doi:10.1021/acs.est.1c03237.

70. Karumanchi, S., Siemiatycki, J., Richardson, L., Hatzopoulou, M. & Lequy, E. Spatial and temporal variability of airborne ultrafine particles in the Greater Montreal area: Results of monitoring campaigns in two seasons. Sci. Total Environ. 771, 144652 (2021).

71. Kerckhoffs, J., Hoek, G., Gehring, U. & Vermeulen, R. Modelling nationwide spatial variation of ultrafine particles based on mobile monitoring. Environ. Int. 154, 106569 (2021).

72. Li, H. Z. et al. Spatially dense air pollutant sampling: Implications of spatial variability on the representativeness of stationary air pollutant monitors. Atmospheric Environ. X 2, 100012 (2019).

73. Patton, A. P. et al. Spatial and temporal differences in traffic-related air pollution in three urban neighborhoods near an interstate highway. Atmos Env. 99, 309–321 (2014).

74. Simon, M. C. et al. Comparisons of traffic-related ultrafine particle number concentrations measured in two urban areas by central, residential, and mobile monitoring. Atmos. Environ. 169, 113–127 (2017).

75. Saha, P. K. et al. Quantifying high-resolution spatial variations and local source impacts of urban ultrafine particle concentrations. Sci. Total Environ. 655, 473–481 (2019).

76. Hudda, N., Simon, M. C., Zamore, W. & Durant, J. L. Aviation-Related Impacts on Ultrafine Particle Number Concentrations Outside and Inside Residences near an Airport. Environ. Sci. Technol. 52, 1765–1772 (2018).

77. Riederer, A. M. et al. Effectiveness of portable HEPA air cleaners on reducing indoor PM _2.5_ and NH _3_ in an agricultural cohort of children with asthma: A randomized intervention trial. Indoor Air ina.12753 (2020) doi:10.1111/ina.12753.

78. Lanphear, B. P. et al. Effects of HEPA Air Cleaners on Unscheduled Asthma Visits and Asthma Symptoms for Children Exposed to Secondhand Tobacco Smoke. Pediatrics 127, 93–101 (2011).

79. James, C. et al. HEPA filtration improves asthma control in children exposed to traffic-related airborne particles. Indoor Air 30, 235–243 (2020).

80. Drieling, R. et al. Randomized Trial of a Portable HEPA Air Cleaner Intervention to Reduce Asthma Morbidity among Latino Children in an Agricultural Community. https://www.researchsquare.com/article/rs-621994/v1 (2021) doi:10.21203/rs.3.rs-621994/v1.

81. Phipatanakul, W. et al. Effect of School Integrated Pest Management or Classroom Air Filter Purifiers on Asthma Symptoms in Students With Active Asthma: A Randomized Clinical Trial. JAMA 326, 839 (2021).

82. Brugge, D. et al. Lessons from in-home air filtration intervention trials to reduce urban ultrafine particle number concentrations. Build. Environ. 126, 266–275 (2017).

